# A systematic review of mask disinfection and reuse for SARS-CoV-2 (through July 10, 2020)

**DOI:** 10.1101/2020.11.11.20229880

**Authors:** Miguel Rothe, Elsa Rohm, Elizabeth Mitchell, Noah Bedrosian, Christine Kelly, Gabrielle String, Daniele Lantagne

## Abstract

We conducted a systematic review of hygiene intervention effectiveness against SARS-CoV-2, including developing inclusion criteria, conducting the search, selecting articles for inclusion, and summarizing included articles. We reviewed 104,735 articles, and 109 articles meeting inclusion criteria were identified, with 33 additional articles identified from reference chaining. Herein, we describe results from 58 mask disinfection and reuse studies, where the majority of data were collected using N95 masks. Please note, no disinfection method consistently removed >3 log of virus irrespective of concentration, contact time, temperature, and humidity. However, results show it is possible to achieve >3 log reduction of SARS-CoV-2 using appropriate concentrations and contact times of chemical (ethanol, hydrogen peroxide, peracetic acid), radiation (PX-UV, UVGI), and thermal (autoclaving, heat) disinfection on N95 masks. N95 mask reuse and failure data indicate that hydrogen peroxide, heat, and UV-GI are promising for mask reuse, peracetic acid and PX-UV need more data, and autoclaving and ethanol lead to mask durability failures. Data on other mask types is limited. We thus recommend focusing guidelines and further research on the use of heat, hydrogen peroxide, and UVGI for N95 mask disinfection/reuse. All of these disinfection options could be investigated for use in LMIC and humanitarian contexts.

**TOC Art:** 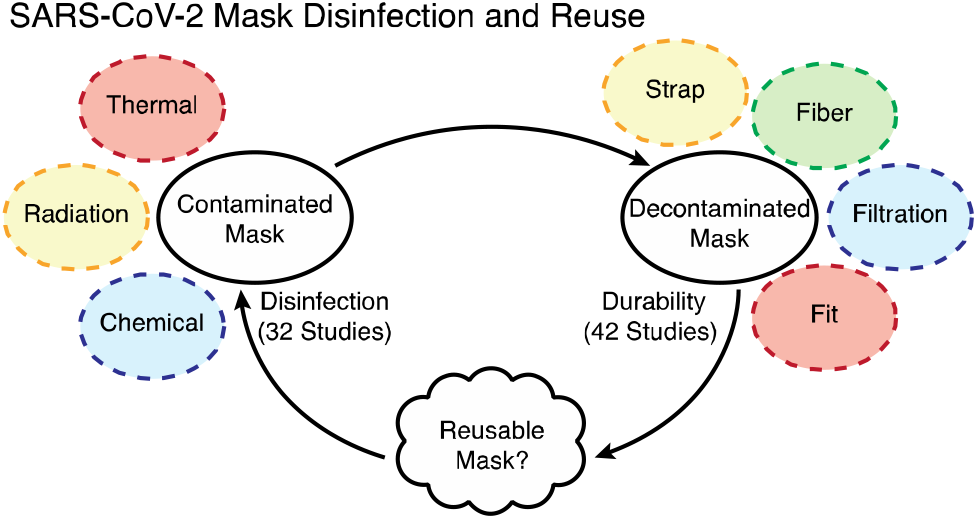

**Synopsis:** In resource-limited contexts, N95s are reused. We recommend using heat, hydrogen peroxide, or UVGI to disinfect and reuse N95 masks.

## Introduction

In December 2019, novel coronavirus SARS-CoV-2 emerged into the human population, leading to a worldwide pandemic.^1^ Due to lack of medical counter-measures, measures to prevent SARS-CoV-2 transmission include physical distancing, masking, hand hygiene, and surface disinfection.^2^

While SARS-CoV-2 has global impact, people living in low- and middle-income countries (LMIC) and humanitarian contexts are particularly impacted by COVID-19.^3^ SARS-CoV-2 transmission is enabled in areas with overcrowded living situations, poor hygiene conditions, and lack of access to personal protective equipment (PPE).^4^ Moreover, refugees, displaced persons, and people in informal settlements in LMIC are particularly vulnerable populations, due to living in crowded conditions with weakened health systems and poor water and sanitation infrastructure.^5, 6^

One hygiene intervention increasingly being used in SARS-CoV-2 contexts is cleaning and/or disinfecting masks for reuse in resource-limited contexts.^7^ Since December 2019, there has been an explosion of research articles related to SARS-CoV-2. The aim of this systematic review was to identify and summarize information on mask disinfection and reuse, in order to develop evidence-based recommendations that are both generally applicable, and specifically applicable to LMIC and humanitarian contexts.

## METHODS

We conducted a previously described systematic review to identify and summarize WASH intervention effectiveness at interrupting SARS-CoV-2 transmission routes.^8^ The review was developed based on the guidelines for the Preferred Reporting Items for Systematic Reviews and Meta-Analyses ^9^ and for mask disinfection and reuse included: 1) a search strategy; 2) inclusion criteria; 3) a selection and data extraction strategy; 4) a framework for appraising risk of bias; and, 5) an analysis plan. Each of these steps is briefly described below (for more information see ^8^).

### Search Strategy

The primary databases searched were the NIH COVID-19 Portfolio (https://icite.od.nih.gov/covid19/help/data-sources) and the CDC COVID-19 Research Articles Downloadable Database (https://www.cdc.gov/library/researchguides/2019novelcoronavirus/researcharticles.html).

The first download was on June 10, 2020, including every article published from database creation on January 22, 2020 until June 10, 2020. The second download occurred on July 10, 2020, and included every article included in the databases from June 10^th^ to July 10^th^, 2020. References were stored in Microsoft Excel (Redmond, WA, USA) and Endnote (Philadelphia, PA, USA), and duplicates were deleted. Reference chaining was completed using reference sections of previous systematic reviews identified.

### Inclusion Criteria

Inclusion criteria were defined according to the populations, interventions, comparisons, outcomes, and study type (PICOS) framework, a model recommended by the Cochrane Library to structure rigorous reviews on health-related questions ^10^. *Populations* included must have been affected by COVID-19. Thus, all age, gender, and socioeconomic populations globally were included. Studies were eligible for inclusion if they included mask disinfection and reuse *interventions*. Specific *comparisons* were not required for inclusion. Studies were eligible for inclusion if they reported *outcome-level results* related to the mask disinfection and reuse. Both published and pre-print *studies* were included if they contained primary data on mask disinfection and reuse. Modeling and prediction studies were not included due to the rapidly changing nature of the COVID-19 pandemic. Only studies in English were included.

### Selection and Data Extraction

Studies were screened by two independent authors in Title, Abstract, and Full Text Screening step for meeting the aforementioned PICOS criteria were excluded. Discrepancies between reviewers were resolved through discussion and consensus.

Relevant data were extracted from each article according to the framework in Waddington *et al* ^11^ including author and publication details, experimental design, and outcomes relevant to mask disinfection and reuse. Data were managed using coding sheets developed in Excel and Google Sheets (Mountainview, CA, USA). Two independent reviewers extracted, discussed, and came to consensus on all data.

### Bias

Because a large number of articles have been published rapidly on SARS-CoV-2 and mask disinfection this review includes both published and pre-print research up to July 10, 2020. Given the data available, it was not possible to systematically assess bias; as such, we divided papers into “published and peer-reviewed” or “pre-print”.

### Analysis

Data were managed and analyzed in Excel and Sheets. All extracted data were tabulated and grouped. Where possible, units were converted to enable data comparability, and missing data were requested from authors.

## RESULTS

From the two downloads of the two databases, a total of 104,735 articles were retrieved. After removal of 8,467 duplicates, 96,268 were title screened. A total of 365 articles passed title and abstract screening, and 109 passed full text screening in addition to 33 articles reference chained from previous systematic reviews. Thus, data were extracted from a total of 142 articles, including 58 articles identified on mask disinfection and reuse.

Of the 58 ^7, 12-68^ mask disinfection and reuse articles, ten were reference chained including nine^59-67^ from three systematic reviews^69-71^, and one^68^ from another article^15^. Of these 58 articles, 30 (52%) were preprints at the time of download^12-16, 23-27, 33, 39-45, 49, 50, 55-63, 66^, and one (2%) was data generated by an FDA contractor with no indication of peer review^68^.

Overall, 16 studies measured disinfection and durability^12, 15, 19, 20, 22, 27-30, 33, 36, 47, 49, 51, 62, 64^, 16 studies measured only disinfection ^13, 18, 23, 24, 31, 38, 40, 41, 43, 44, 48, 56, 63, 65-67^ and 26 studies measured only durability^7, 14, 16, 17, 21, 25, 26, 32, 34, 35, 37, 39, 42, 45, 46, 50, 52-55, 57-61, 68^. Thus, 32 studies measured disinfection and 42 measured durability.

### Mask Disinfection

In total, 92 individual mask disinfection tests were conducted across 32 studies^12, 13, 15, 18-20, 22-24, 27-31, 33, 36, 38, 40, 41, 43, 44, 47-49, 51, 56, 62-67^. The masks tested included: 75 tests with N95s (82%), 5 (5%) with cloth masks, 4 (4%) with surgical masks, three (3%) with KN95s, three (3%) with Tyvek, and 1 (1%) with a plastic face shield and 1 (1%) unspecified. Please note 78 (85%) are considered respirators (N95s and KN95s).

Disinfection methods were grouped into three categories: chemical disinfection, radiation disinfection, and thermal disinfection. In total, 31 chemical disinfection tests were conducted (34% of tests), including 15 tests with various forms of hydrogen peroxide (aerosolized, gas plasma, ionized, vapor), ozone gas (11), ethanol (2), peracetic acid with hydrogen peroxide (1), peracetic acid (1), and ethylene oxide (1). In total, 18 radiation disinfection tests were conducted, including 15 with ultraviolet germicidal irradiation (UVGI) and three with pulsed xenon UV (PX-UV). In total, 43 (47%) thermal disinfection tests were conducted, including moist heat (17 samples), dry heat (14), steaming (5), microwave generated steam (MGS) (5), autoclave (1), and heat (1). Please note disinfection efficacy was calculated using log reductions, or if not available, in a “complete inactivation” binary metric as reported by study authors.

Of the 92 total tests, 21 were conducted with MS-2, 15 with SARS-CoV-2, 12 with IAV, 8 with Phi6, 8 with H1N1, 6 with PCRV, 6 with a mixture (MS2, Phi6, IAV, MHV), 3 with MHV, 3 with H5N1, 2 with P22, and 1 each with Tulane virus, TGEV, Rotavirus, PPV, HCoV-229E, Canine parvovirus, BVDV, and adenovirus.

As can be seen, while the majority of data were collected on N95s, there was high variability in the number of disinfectants tested against SARS-CoV-2 and surrogates used for testing. Limiting the results to N95 masks only, there were 15 tests using SARS-CoV-2, 18 using MS2, eight using Phi6, eight using H1N1, and six using IAV.

In the 15 SARS-CoV-2 N95 mask samples, eight were chemical (hydrogen peroxide^15, 29, 30, 36, 48^, ethanol^15, 29^, peracetic acid^30^), four were radiation (PX-UV^13^, UVGI ^15, 29, 31^), and three were thermal (autoclave^30^, dry heat^29, 49^). All applications of hydrogen peroxide (10-210 minute exposure), ethanol (spraying, saturation), and 10% peracetic acid (60 minute exposure) achieved SARS-CoV-2 inactivation or >3 log reduction. PX-UV and UVGI achieved >3 log reduction with ≥5 minute contact time. Dry 70°C heat for 50 minutes and autoclaving at 121°C for 15 minutes led to >5 and >6 log reduction, respectively. Please note all methods tested achieved inactivation (binary) or >3 log reduction except for UVGI at 2 minutes contact time.

In the 18 MS2 results^22, 23, 44, 56, 64, 66, 72^, UVGI for 1-10 minutes did not achieve >3 log reduction, nor did 70°C dry heat for 15-30 minutes. Steam for 10 seconds - 15 minutes did achieve >3 log reduction. Moist heat at 72-82°C for 30 minutes did achieve >6 log reductions when humidity was >25% or >50%. The results from the eight H1N1 results^33, 38, 63, 65^ were presented in log reduction ranges, not specific log reductions, which precludes analysis herein. In the eight Phi6 results^27, 40, 56, 72^, moist heat at 72-82°C for 30 minutes achieved <6 or >6 log reduction but inaccurate humidity readings occlude results, 7% aerosolized hydrogen peroxide had >6 log reductions with 30 minute exposure, and VHP at 16 gram/minute had >4 log after 30-40 minute exposure time; please note UVGI for one minute had only 1.5 log reduction. In the six IAV samples^41, 43, 56, 63^, ozone removed 1-2 log after 40 minute exposure to 20 ppm, and moist heat and MGS achieved >3 log reduction after 30 minutes exposure to 60-82C moist heat for 30 minutes and 2 minute exposure to MGS.

Five tests were conducted on cloth masks^23, 41^, using ozone gas against IAV, and heat/steam against MS2. Steaming for 15 minutes achieved >15 log reduction (the only method to achieve >3). Four tests were conducted on surgical masks^24, 64^, using UVGI for 2 minutes, dry heat at 102°C for 60 minutes, VHP for 20 minutes, and MGS for 30 minutes; all achieved >3 log reduction. In three tests on Tyvek^41^, ozone gas at 20 ppm for 40 minutes led to ∼1-2 log reductions of IAV. On one face shield^67^, PX-UV reduced canine parvovirus by >4 log with 5 minute exposure time. Studies noted radiation disinfection did not deactivate viruses in a uniform manner due to the complex shapes of masks, thus masks should not be stacked if UV disinfection is used.

Please note no disinfection method consistently removed >3 log reduction of virus irrespective of concentration, contact time, temperature, and humidity. However, results did show that it is possible to achieve >3 log reduction of SARS-CoV-2 using appropriate concentrations and contact times of chemical (ethanol, hydrogen peroxide, peracetic acid), radiation (PX-UV, UVGI), and thermal (autoclaving, heat) disinfection.

### Mask reuse

Mask durability was assessed based on four criteria: filtration, fit, fiber resilience, and strap performance. Disinfection cycles were also noted, and the maximum amount of cycles was determined when a failure was recorded in any of the four criteria.

In total, 159 individual mask reuse tests were conducted in 42 studies ^7, 12, 14-17, 19-22, 25-30, 32-37, 39, 42, 45-47, 49-55, 57-62, 64, 68^. The masks tested included: 114 tests with N95s (72%), an additional four tests with KN95s (2.5%), seven tests with folded or molded or HKYZ N95s (4.4%), four tests with FFPs (2.5%), and two tests with respirator fabric (1%). Thus a total of 131 tests (82%) were conducted with respirators. Additionally, 11 tests were conducted with surgical masks (7%), five with nanofiber filter masks (3.1%), four with procedure masks (2.5%), three with sterilization wrap (2%) and EX101 masks (2%), and one (1%) with a cloth mask, and one (1%) unspecified.

Disinfection methods were grouped into three categories: chemical disinfection, radiation disinfection, and thermal disinfection. In total, 65 chemical reuse tests were conducted (41% of tests), including 20 tests with forms of hydrogen peroxide, ethanol (20), isopropyl alcohol (7), bleach (5), ethylene oxide (5), soap and water (3), ozone gas (3), peracetic acid (1), and household detergent (1). In total, 24 radiation reuse tests were conducted (15%), including 15 with ultraviolet germicidal irradiation (UVGI), five with gamma radiation, three with microwaves, and one with pulsed xenon UV (PX-UV). In total, 70 (44%) thermal reuse tests were conducted, including dry heat (23 samples), autoclave (16), steam (10), moist heat (9), microwave generated steam (6), heat (3), hot water soak (2), and boiling (1).

The majority of data were collected on N95s. Given the wide differences in test conditions, results are summarized by disinfection agent for N95s only, for agents with >5 samples or those agents identified in the disinfection efficacy section as achieving >3 log reduction of SARS-CoV-2, including ethanol, hydrogen peroxide, peracetic acid, PX-UV, UVGI, autoclaving, and heat.

Autoclaving (n=14)^14, 30, 34, 42, 45, 55, 57, 59^ and ethanol disinfection (n=14)^15-17, 29, 42, 45, 53, 58^ led to failures within two disinfection cycles in 12 and 13 tests, respectively. This indicates these methods cannot be utilized for N95 reuse.

Steam (n=5)^16, 26, 42, 52^ and moist heat (n=9)^7, 19, 28, 32, 45, 61, 62^ had mixed results, with 3 failures in 1-5 cycles (but two tests that lasted up to 1-10 cycles), 4 failures in 0-4 cycles (and four tests with no failure in 1-10 cycles and one not recorded), respectively. More research is indicated to understand these results. Hydrogen peroxide disinfection (n=20)^15, 19, 27, 29, 30, 36, 45, 51, 57, 59-61, 68^, dry heat (n=19)^12, 16, 19, 21, 29, 33-35, 42, 45, 49, 50, 52, 54^, UVGI (n=13)^15, 16, 29, 35, 42, 45, 46, 52, 59-62^, and ethylene oxide (n=5)^19, 59-61^ all had promising results. With hydrogen peroxide, no failures were seen in the course of the study in 14 tests (1-14 cycles), two failed at 14 and 29 cycles, and – using HPGP and HPAH, there were failures at 1-4 and 9 cycles, respectively. In dry heat tests, 15 masks lasted the maximum number of times tested (1-20 cycles), and four failed with 0-3 disinfection cycles. In UVGI tests, 10 masks lasted the number of times tested (1-20 cycles), and three failed within 2 disinfection cycles. In ethylene oxide tests, none failed within the testing (1-3 cycles). Please note peracetic acid and PX-UV had one test each, with no failures in the 10 cycles tested.

### Summary

Our overall goal was to complete a systematic review of transmission pathways of SARS-CoV-2 that could be interrupted with WASH interventions, with a focus on LMIC and humanitarian contexts. After completing title, abstract, and full-text review of 104,735 articles, 109 articles meeting inclusion criteria were identified, with 33 additional articles identified from reference chaining. Information on surface disinfection has been published separately^8^; herein we described results from 58 mask disinfection and reuse summaries.

We identified a large amount of data on mask disinfection and reuse, although much was not comparable as many studies lacked specificity on methods, and tested a wide range of conditions on a small number of samples. This led to data from multiple articles summarized to generate themes. Overall, when comparing the list of efficacious disinfectants (ethanol, hydrogen peroxide, peracetic acid, PX-UV, UVGI, autoclaving, and heat) with the reuse information (hydrogen peroxide, heat, and UV-GI promising, peracetic acid and PX-UV need more information, and autoclaving and ethanol lead to failures), it is recommended to focus guidelines and future research on the use of heat, hydrogen peroxide, and UVGI. For radiation, non-uniform distribution across N95 masks was noted, when can be managed by rotation and ensuring no shadowing by straps/other masks^16^. All of these options could be investigated for use in LMIC and humanitarian contexts. Further research is recommended on non-N95 masks, as there is a lack of research on cloth masks in particular.

Limitations to this work include that pre-prints were included in review, bias was not assessed beyond noting pre-print percentage, and after completing the review we (based on information available) determined to focus this sub-paper on mask disinfection and reuse. Additionally, there is likely more relevant data published after the final download date of July 10, 2020. We plan to update this review for a second publication in early-mid 2021. We do not feel these limitations impact the results presented herein. Please note all extracted data is available in an Excel file in Supplementary Information for open-access use.

In summary, while SARS-CoV-2 has had a global impact, people living in LMIC are disproportionately impacted due to inability to adapt recommended safety measures, lack of resources, and underlying health conditions. We identified that no disinfection method consistently removed >3 log reduction of SARS-CoV-2 virus irrespective of concentration, contact time, temperature, and humidity. However, results did show that it is possible to achieve >3 log reduction of SARS-CoV-2 using appropriate concentrations and contact times of chemical (ethanol, hydrogen peroxide, peracetic acid), radiation (PX-UV, UVGI), and thermal (autoclaving, heat) disinfection, and that hydrogen peroxide, heat, and UV-GI have promise to allow mask reuse without mask failures. Further research on mask disinfection and reuse – on a range of masks – is needed to develop evidence-based recommendations to protect people by ensuring safety of PPE in contexts where masks are re-used.

## Data Availability

All data is appended in the attached Annex.

## Acknowledgements

The authors would like to thank the funders of this work, including: TISCH College of Civic Life at Tufts University (funding for Bedrosian, Rohm, Rothe), D. Lantagne discretionary funding (funding for Bedrosian, Mitchell, Rohm, Rothe), and NSF 1657218 (funding for Kelly).

## SUPPLEMENTAL INFORMATION

Table S1. Mask disinfection and reuse table

